# Changes in self-reported health and wellbeing outcomes in 36,951 primary school children from 2014 to 2022 in Wales: A descriptive analysis using annual survey data

**DOI:** 10.1101/2023.06.15.23291464

**Authors:** Johanna Einhorn, Michaela James, Natasha Kennedy, Emily Marchant, Sinead Brophy

**Affiliations:** National Centre for Population Health and Wellbeing Research, Data Science Building, Swansea University, Singleton Park, Swansea, SA2 8PP; Department of Education and Childhood Studies, School of Social Sciences, Swansea University, Singleton Park, Swansea, SA2 8PP

## Abstract

This study examines the changes in childhood self-reported health and wellbeing between 2014 and 2022. An annual survey delivered by HAPPEN-Wales, in collaboration with 500 primary schools, captured self-reported data on physical health, dietary habits, mental health, and overall wellbeing for children aged 8-11 years. The findings reveal a decline in physical health between 2014 and 2022, as evidenced by reduced abilities in swimming and cycling. For example, 68% of children (95%CI: 67%-69%) reported being able to swim 25m in 2022, compared to 85% (95% CI: 83%-87%) in 2018. Additionally, unhealthy eating habits, such as decreased fruit and vegetable consumption and increased consumption of sugary snacks, have become more prevalent. Mental health issues, including emotional and behavioural difficulties, have also increased, with emotional difficulties affecting 13%-15% of children in 2017-2018 and now impacting 29% of children in 2021-2022. Moreover, indicators of wellbeing, autonomy, and competence have declined. Importantly, this trend of declining health and wellbeing predates the onset of the Covid-19 pandemic, suggesting that it is not solely attributed to the pandemic’s effects. The health of primary school children has been on a declining trajectory since 2018/2019 and has continued to decline through the COVID recovery period. The study suggests that these trends are unlikely to improve without targeted intervention and policy focus.

## Introduction

The foundation of adulthood is laid in childhood, where experiences shape life achievements, mental health, and overall wellbeing [1]. It is noted that 50% of mental health problems are established by the age of 14 [2], and stressors in childhood can influence brain development [3]. Thus, the physical and mental health, as well as the overall wellbeing and self-confidence, of the nation’s school-aged children is vital to the health of the next generation.

The incidence of mental health disorders in childhood is increasing; global research indicates that mental health problems such as anxiety, depression, and behavioural problems increased among children between 2014 to 2020 [4]. Observations of data from the United Kingdom (UK) revealed that mental health conditions rose from 1 in 9 to 1 in 6 between 2017 to 2021 [5].

Moreover, trends observed in England and Wales have reported a decline in wellbeing, especially as children age and particularly in girls [6–8]. In addition, recent evidence indicates that children’s physical health is deteriorating; English children were demonstrated to have an increase in Body Mass Index (BMI) and decrease in general fitness in 2020 [9], resulting in an increase in obesity rates since 2017 [8].

Legislative measures, such as the Wellbeing of Future Generations Act (WFGA) [10] in Wales, as well as policies concerning physical activity and play (e.g., Children and Families (Wales) Measure 2010, and the Play Sufficiency Duty [11]), have raised awareness of the need to improve the health and wellbeing of young people. The Covid-19 pandemic has potentially contributed to a decline in health and wellbeing, as access to socialisation, support, and physical activity has been reduced due to school closures aimed at preventing virus transmission [12–14]. Persistently elevated levels of mental health disorders in 2021 indicate that the full impact of the pandemic remains unaddressed or that overall health outcomes are trending downward [8].

Furthermore, amidst the Covid-19 pandemic, children have been exposed to a risk of contracting and transmitting the virus, as well as experiencing the associated health consequences. They have also faced elevated risks of reduced access to medical care, poverty, and social isolation due to Covid-19 restrictions [15]. These challenges have arisen in conjunction with the UK’s departure from the European Union [16] and the current “cost of living crisis” [17, 18], subsequently presenting significant obstacles for children and society’s support systems.

This research aims to explore childhood trends between 2014 and 2022; examining changes in childhood experiences and how growing up during periods of political upheaval and crisis (e.g. the Covid-19 pandemic) has affected the health and wellbeing of children. This study employs data from a self-report survey administered by HAPPEN (Health and Attainment of Pupils in Primary EducatioN)-Wales to 36,951 children aged 8-11 across Wales. The survey covers aspects such as general health, physical activity, sleep, dietary habits, wellbeing, social contacts, mental health, and their experience of growing up.

## Materials and Methods

### Study Design

This study employs data collected by HAPPEN-Wales between 2014 and 2022. HAPPEN-Wales was officially established at Swansea University in 2015 and has been co-produced alongside school senior leaders, teachers, and pupils. Prior to this HAPPEN data was collected via ‘Fitness Fun Days’. Senior leaders advocated for collaboration and a joined-up approach to prioritising health and wellbeing within the school setting [19] to bring together education, health, and research in line with the *Curriculum for Wales* proposals for health and wellbeing Wales, implemented in primary schools from September 2022.

Participating schools engage with the initiative by completing an online self-report survey with their pupils aged 8 - 11 years (years 4, 5 and 6). The survey encompasses various dimensions of health and wellbeing, including nutrition, physical activity, sleep, mental health, and concentration [20]. The survey is included as supplementary file 1. The modified version circulated during Covid-19 can be observed in supplementary file 2. The latest version of the survey was granted ethical approval in November 2022 by Swansea University’s Medical School (2017-0033I).

### Participants

HAPPEN is a pan-Wales infrastructure that delivers an online health and wellbeing survey to schools. To recruit, all primary schools in Wales are contacted via direct email, social media campaigns (including paid advertisements on Facebook and Twitter), and promotion through key stakeholders (e.g., regional education consortia) throughout the academic year. Schools that agree to participate are provided with information sheets to be shared with parents/guardians, who are given the opportunity to opt their child out of the survey. This opt-out approach to recruiting participants was implemented to establish a representative sample that encompasses the diverse population of children across Wales. Child information sheets are distributed, and assent is obtained at the start of the survey. Typically, the survey is completed in school as part of a lesson, supervised by a teacher. The exception for this is during 2020, where the survey was adapted and completed at home during Covid-19 enforced school closures [14]. Prior to 2018, data were collected in South Wales specifically before HAPPEN expanded pan-Wales from 2018.

### Data Collection

The data were collected between September 2014 and December 2022. All children in Wales aged 8 - 11 are eligible to participate in the survey. The process of data coding involved four researchers. The first researcher (MJ/EM) downloaded the raw data, cleaned the data, and generated a unique participant ID number before removing identifiable information. This process protects participants’ anonymity. Raw data was coded using STATA (version 16) [21] to produce a dataset for the purpose of analyses. A second researcher (JE) conducted analysis of the data with a third (NK) validating the initial analysis.

### Analysis

For data analysis and statistics procedures, the R software (version 4.2.2) [22] was used, employing the packages “*lubridate”*, “*MASS”,* and “*tidy verse”.* From the survey, the variable “date participated” was extracted for use as the explanatory variable. The examined variables encompassed a range of factors, including the quantity of fruit and vegetables consumed, frequency of fatigue experienced per week, duration of screen time per week, proficiency in riding a bike without stabilisers, ability to swim 25 metres unassisted, self-reported happiness level with friends on a scale of 1 to 10, self-perceived competence level, level of worry experienced, emotional and behavioural difficulty scores derived from the Me and My Feelings questionnaire, self-perceived safety of the child’s area on a scale of 1 to 10, agreement on the importance of physical activity, number of friends seen in the past week, and presence of cold symptoms within the previous week. Additionally, demographic variables such as “age”, “gender”, “WIMD2019Quartile”, and “ChildIndex2011Quintile” were included as variables and covariates (supplementary file 3).

The dependent variables were sorted into four categories. The first category describes variables with binary responses (yes or no). The second category encompassed variables with a concrete numerical answer such as “*How many portions of fruits or vegetables did you eat yesterday*”.

The third category consisted of variables with a categorical scale ranging from 0 to 4, where each category represented a distinct response. The final category involved variables with a scale of one to 10. Only complete data were included in the analysis.

For variables from category one, the percentage of children answering ‘yes’ per month and year was calculated. For variables from the second, third, and fourth category the percentage of children choosing a specific category was determined on a monthly and yearly basis. In cases where “*I do not know*” or “*I am not sure*” were selected as answers, these responses were treated as missing data rather than separate categories due to the small number of cases. Additionally, for all results 95% confidence intervals were calculated.

Regression analysis adjusting for age, gender and deprivation was used to examine change in health before and after 2020 (as a time marker of the Covid-19 pandemic) using interrupted time series.

## Results

This study uses data from a total of 36,951 children from 2014 to 2022. Of this number, 47% were boys, 49% were girls and 3% preferred not to say. The average age of children was 9.35 years. In terms of deprivation levels, 15% of participants were classified in the most deprived quartile (1), 12% in quartile 2, 14% in quartile 3, and 7% in quartile 4 (least deprived). 38% of participants did not have deprivation associated data. Overall findings can be observed in table 1.

**Table 1.** Overall Findings In Children’s Self-Reported Health and Wellbeing from 2014 to 2022.

|  |  |  |  |  |  |  |  |  |  |
| --- | --- | --- | --- | --- | --- | --- | --- | --- | --- |
| VARIABLE | 2014 | 2015 | 2016 | 2017 | 2018 | 2019 | 2020 | 2021 | 2022 |
| Fruit & Veg<br>*Percentage eating one or two portions per day* | 29% | 21% | 24% | 18% | 22% | 33% | 29% | 28% | 29% |
| CI | 0.24-0.33 | 0.17-0.23 | 0.22-0.26 | 0.15-0.2 | 0.19-0.25 | 0.32-0.34 | 0.28-0.30 | 0.28-0.30 | 0.28-0.30 |
| Sugary Snacks<br>*Percentage eating more than two sugary snacks a day* | 35% | 30% | 30% | 27% | 28% | 34% | 39% | 40% | 37% |
| CI | 0.30-0.40 | 0.27-0.33 | 0.28-0.32 | 0.24-0.30 | 0.25-0.31 | 0.33-0.35 | 0.38-0.40 | 0.39-0.41 | 0.36-0.38 |
| Feeling Tired<br>*Percentage tired more than three days a week* | 41% | 35% | 38% | 39% | 40% | 48% | 47% | 51% | 54% |
| CI | 0.36-0.46 | 0.32-0.39 | 0.36-0.41 | 0.36-0.42 | 0.37-0.43 | 0.46-0.49 | 0.46-0.48 | 0.50-0.52 | 0.52-0.55 |
| Ride A Bike<br>*Percentage of children able to ride a bike* | 90% | 90% | 92% | 90% | 91% | 87% | 86% | 86% | 87% |
| CI | 0.88-0.94 | 0.89-0.93 | 0.91-0.93 | 0.88-0.92 | 0.88-0.92 | 0.86-0.87 | 0.85-0.86 | 0.86-0.87 | 0.84-0.87 |
| Swim 25m<br>*Percentage of children able to swim 25m* | 72% | 78% | 80% | 81% | 85% | 74% | 72% | 71% | 68% |
| CI | 67-77 | 75-81 | 79-82 | 0.79-0.84 | 0.83-0.87 | 0.73-0.75 | 0.71-0.73 | 0.70-0.72 | 0.67-0.69 |
| Feeling Safe<br>*Percentage feeling more than 5/10 safe* |  |  |  | 61% | 87% | 81% | 84% | 84% | 83% |
| CI |  |  |  | 0.57-0.66 | 0.85-0.89 | 0.80-0.82 | 0.83-0.84 | 0.83-0.85 | 0.82-0.84 |
| Happiness with Friends<br>*Percentage of children reporting more than 5/10* |  |  |  | 88% | 94% | 89% | 92% | 92% | 91% |
| CI |  |  |  | 0.86-0.90 | 0.93-0.96 | 0.89-0.91 | 0.92-0.93 | 0.92-0.93 | 0.91-0.92 |
| General Competency<br>*Percentage of children agreeing* |  |  |  | 92% | 94% | 89% | 88% | 89% | 88% |
| CI |  |  |  | 0.90-0.94 | 0.92-0.97 | 0.88-0.89 | 0.87-0.89 | 0.88-0.89 | 0.87-0.89 |
| Worry A Lot<br>*Percentage of children that worry a lot* |  |  |  | 12% | 10% | 17% | 16% | 20% | 20% |
| CI |  |  |  | 0.10-0.13 | 0.08-0.12 | 0.16-0.18 | 0.15-0.17 | 0.19-21 | 0.17-0.20 |
| Emotional Difficulties<br>*Percentage reporting clinical emotional difficulties.* |  |  |  | 13% | 15% | 23% | 24% | 29% | 29% |
| CI |  |  |  | 0.11-0.15 | 0.12-0.17 | 0.22-0.24 | 0.23-0.25 | 0.28-0.30 | 0.27-0.30 |
| Behavioural Difficulties<br>*Percentage reporting clinical behavioural difficulties.* |  |  |  | 18% | 17% | 25% | 24% | 24% | 26% |
| CI |  |  |  | 0.16-0.20 | 0.14-0.19 | 0.24-0.26 | 0.23-0.25 | 0.23-0.25 | 0.25-0.27 |
| Feel Lonely<br>*Percentage of children feeling lonely sometimes or a lot* |  |  |  | 55% | 53% | 60% | 61% | 64% | 63% |
| CI |  |  |  | 0.52-0.58 | 0.49-0.56 | 0.59-0.61 | 0.60-0.62 | 0.63-0.65 | 0.61-0.64 |
| Out of School Clubs<br>*Percentage of children in more than two clubs* |  | 35% | 32% | 40% | 44% | 37% | 29% | 33% | 32% |
| CI |  | 0.32-0.38 | 0.30-0.35 | 0.67-0.43 | 0.41-0.48 | 0.36-0.38 | 0.28-0.30 | 0.32-0.34 | 0.30-0.33 |
| General Wellbeing<br>*Percentage of children reporting higher than 6/10* |  |  |  | 85% | 91% | 86% | 88% | 88% | 87% |
| CI |  |  |  | 0.83-0.87 | 0.90-0.93 | 0.85-0.86 | 0.87-0.88 | 0.88-0.89 | 0.87-0.88 |
| General Autonomy<br>*Percentage of children agreeing they have autonomy* |  |  |  | 91% | 95% | 92% | 92% | 92% | 92% |
| CI |  |  |  | 0.87-0.93 | 0.93-0.97 | 0.91-0.93 | 0.92-0.93 | 0.91-0.93 | 0.91-0.92 |

### Physical health

For this study, physical health includes physical competency measures of the ability to swim, ability to ride a bike, understanding of the benefits of physical activity, diet, and tiredness (table 1). The ability to swim was measured by the percentage of children self-reporting they were able to swim 25m. This decreased rapidly in the last few years and has not recovered (Fig1). Currently, only 68% (95% CI: 67-69%) of children reported to be able to swim 25m in 2022, compared to 85% (95% CI: 83%-87%) in 2018.

**Fig1.**
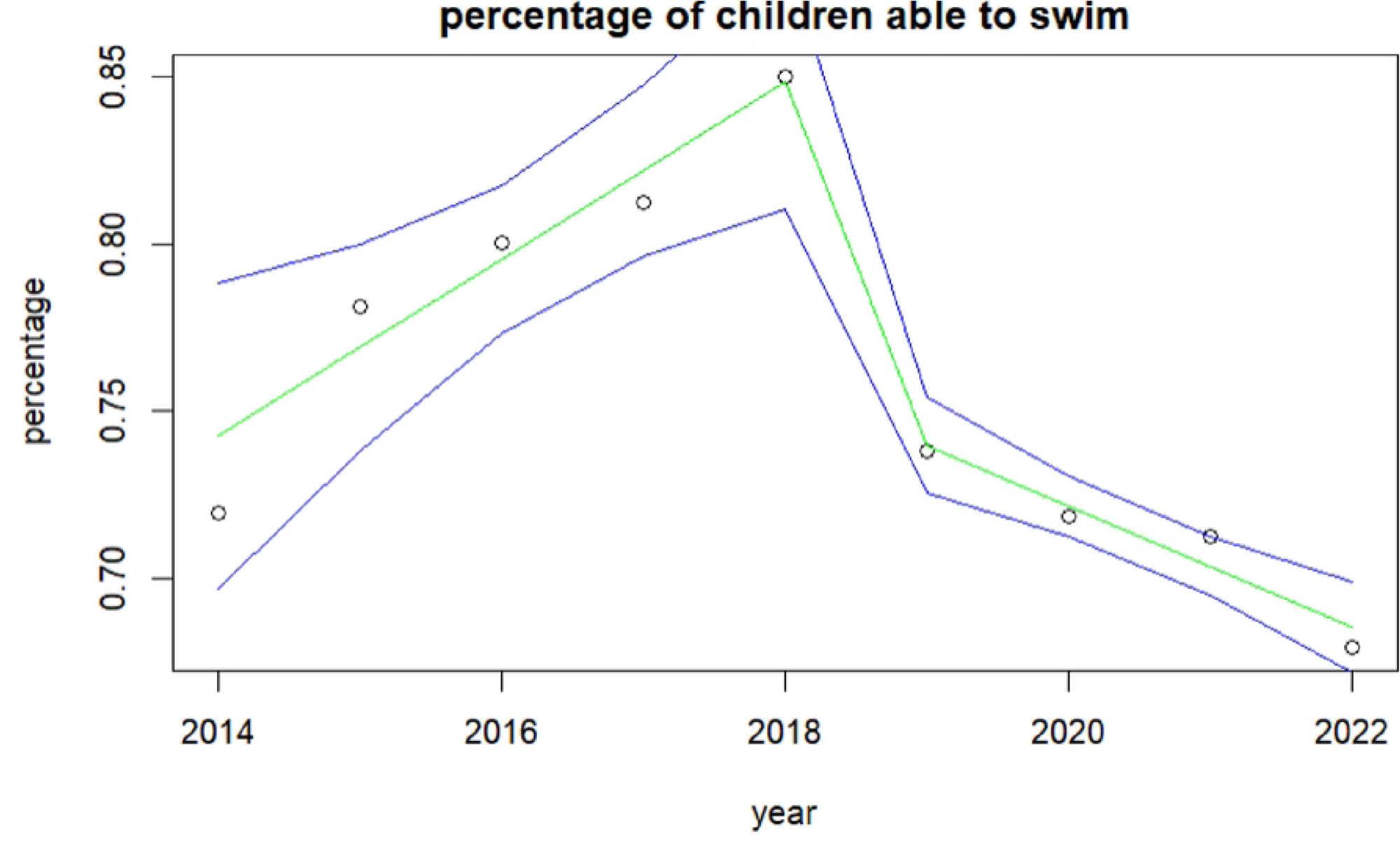
Children’s ability to swim since 2014.

This analysis also shows an association between the ability to ride a bike and the number of friends children are having over their houses with those with more friends more likely to be able to ride a bike. As well as this, an association is observed between the ability to ride a bike and tiredness; higher tiredness was reported in those who cannot ride a bike.

The data shows an overall increase of tiredness among all children (Fig2). From 41% of children feeling tired more than 3 days a week in 2014 (95%CI: 36%-46%) this is relatively unchanged until 2019 where the mean increases to 51% (95%CI: 50%-52%) and 54% (95%CI: 53%-55%) in 2022. ANOVA analysis shows an interaction between tiredness and the number of sugary snacks consumed with increases in tiredness seen as sugary snack consumption increases.

**Fig2.**
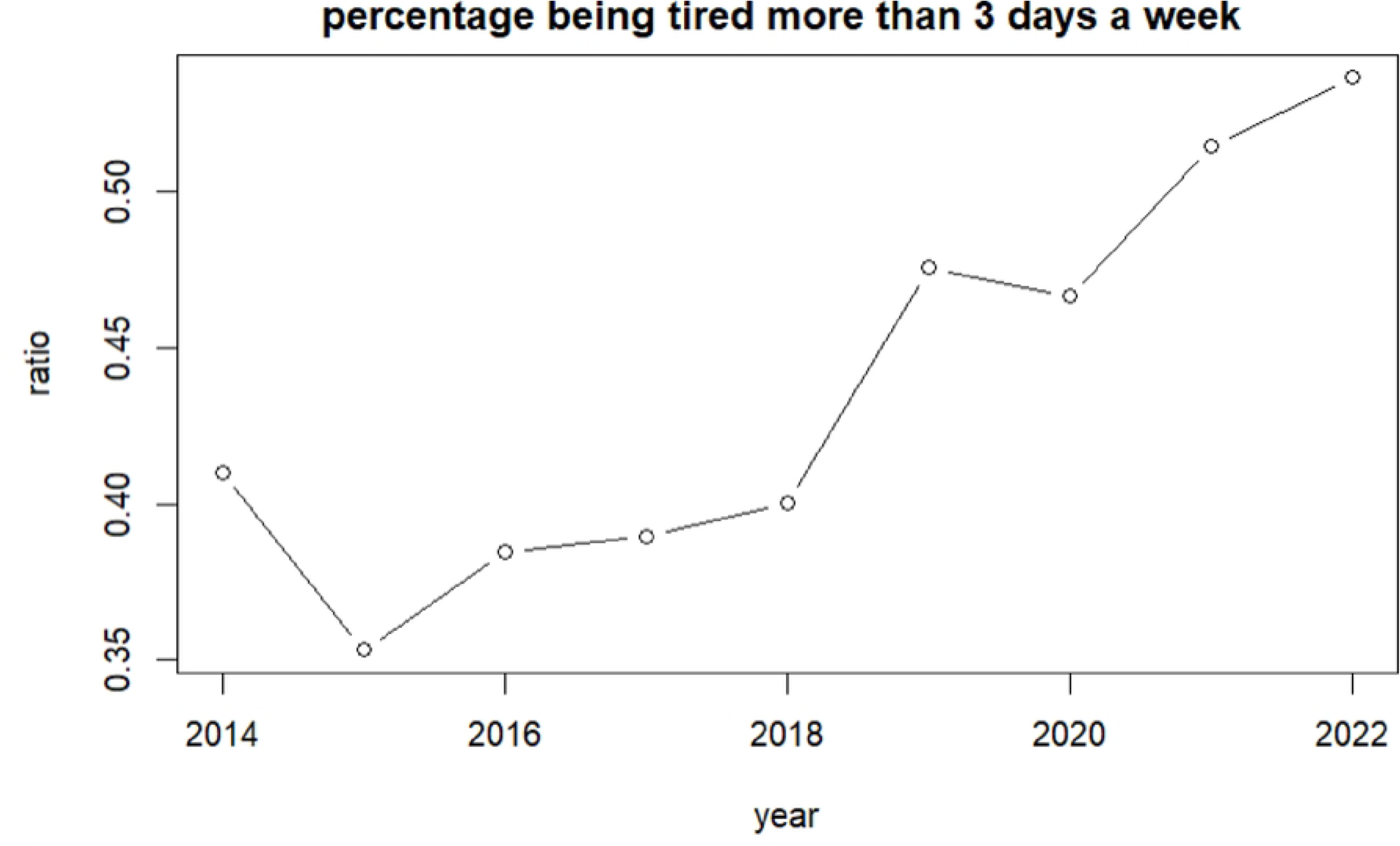
Children’s self-reported tiredness since 2014.

The average number of sugary snacks children consumed per day increased rapidly in 2020 but was already increasing from 28% (95%CI: 25%-32%) of children eating more than two sugary snacks in 2018 to 34% (95%CI: 33%-35%) in 2019. In 2020, it went up to 39% (95%CI: 38%-40%) and has remained at this level. Regression analysis shows a significant increasing trend.

### Mental Health and Wellbeing

The percentage of children reporting emotional difficulties increased from 13% (95%CI: 11%-15%) in 2017 to 23% (95%CI: 23%-25%) in 2019 and to 29% (95%CI: 27%-30%) in 2022 (Fig3). Higher values represent higher difficulties. ANOVA analysis indicates differences between genders with boys reporting lower emotional difficulties than girls. The highest difficulties are children who preferred not to state their gender.

**Fig3.**
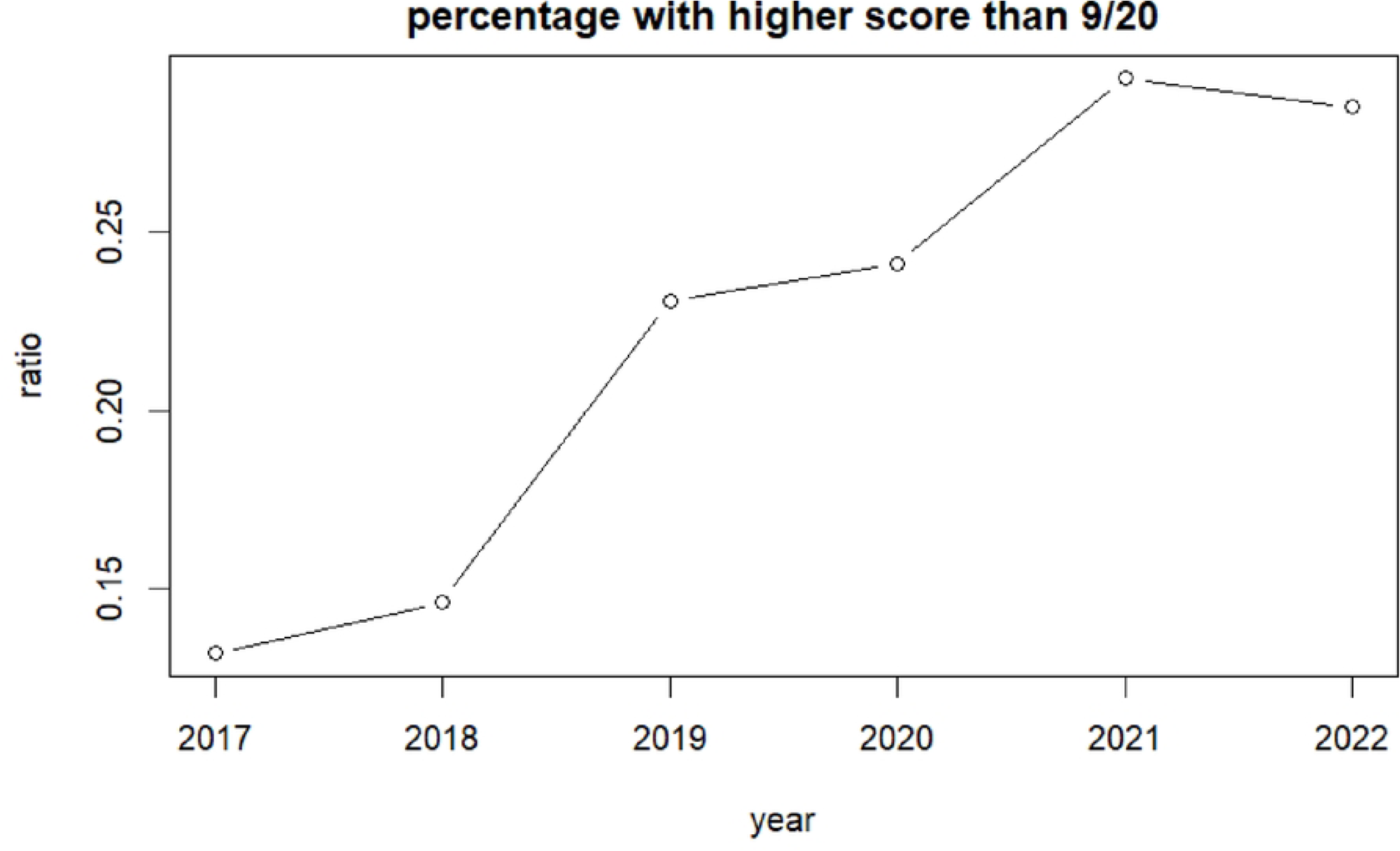
Increase in emotional difficulties since 2014.

Behavioural difficulties increased in general, especially in girls. The percentage of children reporting behavioural difficulties increased from 18% (95%CI: 16%-20%) in 2017 to 25% (95%CI: 24%-26%) in 2019 (Fig4).

**Fig4.**
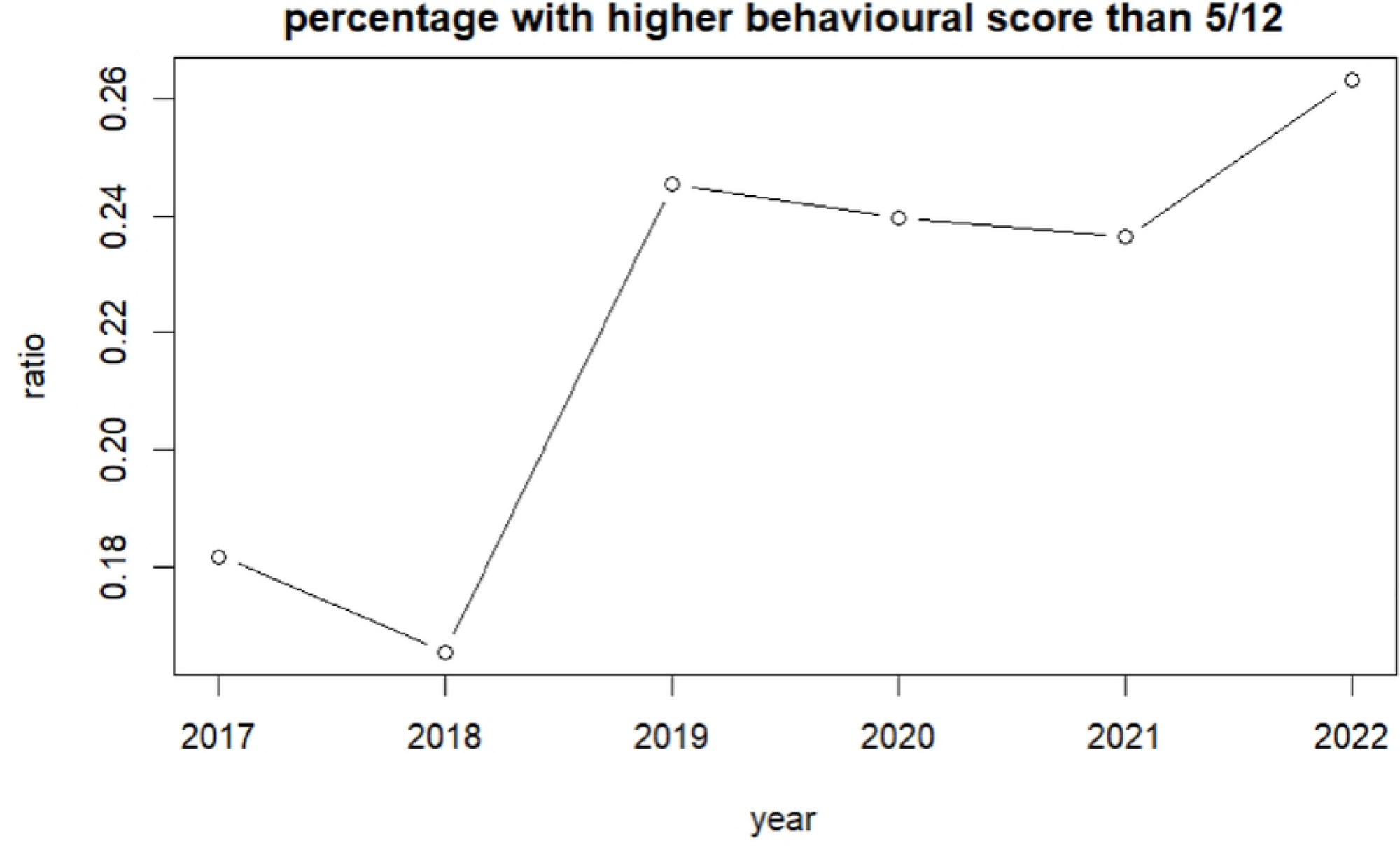
Increase in behavioural difficulties since 2014.

Since 2017 children reporting they worry a lot has increased from 17% (95%CI: 16%-18%) in 2017 to 20% (95%CI: 17%-20%) in 2022. Regression analysis shows a significant inclining trend.

### Socialisation

Since 2019, the number of children feeling lonely increased from 55% (95%CI: 52%-58%) to 63% (95%CI: 61%-64%) in 2022. Additionally, ANOVA analysis shows an interaction between increased loneliness and a higher number of sugary snacks consumed.

## Discussion

The aim of this research was to explore childhood trends over the last eight years; examining the changes in childhood health and wellbeing. The health and wellbeing of children has been declining globally and data has indicated that mental health conditions are rising. The Covid-19 pandemic and its associated restrictions have potentially impacted this further. This, alongside points of political and societal upheaval in the UK (e.g., Brexit and the ‘cost of living’ crisis) have presented challenges for society.

This study has identified some key findings in children’s health and wellbeing from 2014 to 2022, with a notable emphasis on the significant declines observed across several areas. Health and wellbeing have been declining for some time, even prior to Covid-19, suggesting that the pandemic has not been the cause of poor health and wellbeing, but has contributed further to the decline.

### Physical Health

The physical health of children has exhibited no improvement since 2014, accompanied by a decline in reported swimming and cycling abilities. These physical skills are essential for developing physical literacy and fundamental movement skills [23], these are often developed during early childhood years. Swimming is often promoted during early years as an avenue of teaching and developing these skills [24]. However, funding cuts in Wales in 2019, which forced changes to swimming provision for children [25], and the subsequent closure of swimming facilities during the pandemic to prevent transmission, have exacerbated this issue. Notably, self-reported swimming ability significantly declined the year prior to the pandemic, highlighting the role of funding redirection and the subsequent lack of accessibility on children’s perception of their physical literacy. Moreover, deprived children are more likely to report not being able to swim, further demonstrating the potential role of the funding cuts in widening inequalities.

Dietary habits have also not improved over time, with no improvements observed in sugar, fruit, and vegetable consumption. Sugar consumption increased rapidly in 2020, coinciding with the Covid-19 pandemic, which caused an increased time at spent home rather than attending school, potentially encouraging less-favourable diets with interruptions to regular, enforced mealtimes during the school day. These observations have been reflected in other studies [14, 30]. Fruit and vegetable intake has also not improved over time, with nearly a third of children in this study reporting eating zero or one portion a day. Other studies noted that more deprived children ate fewer portions of fruit and vegetables during the pandemic [14], and free school meals, a key policy that helps reduce dietary inequalities, remained inaccessible to almost half of all children on free school meals during pandemic restrictions [31].

The combination of fewer physical skills, a lack of understanding of the benefits of physical activity, and a rise in sugary snack consumption could lead to an increase in sedentary behaviour, obesity, and related negative health behaviours among this generation of children, potentially increasing the risk of a range of non-communicable diseases such as diabetes. It is important to note that all the above variables were already showing a decreasing trend in 2019 before the pandemic, indicating that the pandemic is not the only risk factor to children’s health and wellbeing. Therefore, the assumption that the situation will naturally recover with restrictions easing cannot be made.

### Mental Health and Wellbeing

The mental health of children has not improved over time and displays a concerning trend. These findings are consistent with previous studies [6–8], and indicate an alarming picture of mental health and wellbeing among school-aged children. Effective and sustainable intervention is necessary to interrupt this trend, particularly for girls who exhibit higher rates of emotional and behavioral difficulties compared to boys. Research indicates that girls are more likely to experience difficulties than boys and this gender gap increases with age [32], underscoring the importance of addressing these trends.

Although this study highlights that this trend was apparent prior to the pandemic, it is possible that periods of isolation, restrictions, and interruption to normal routine have highlighted and exacerbated these issues. Research indicates that a proportion of young people reported the pandemic had worsened their mental health [33]. The pandemic may have exacerbated these trends by increasing worry and uncertainty over young people’s futures, particularly in terms of schooling, where young people reported difficulties with online learning [34].

However, these trends were reported among young people in secondary school and higher education. These trends have been ongoing since 2019, where the most significant declines were observed in this study. It is plausible that younger children were happier and healthier during lockdowns, especially as this study notes some increases in wellbeing in 2020. It is critical that children of all ages are heard, with their wants and needs recorded and addressed, as they differ by age and gender. During the pandemic, younger children valued the ability to play with their friends, while older children were concerned about future prospects and online learning [34]. Addressing areas of support tailored to their specific needs is necessary to alleviate negative trends.

Friendships are vital to children’s social lives, providing them with early experiences of relationships and support beyond their families [35]. This research suggests that children are not socializing with friends as much as they used to and feel lonelier, which may have been exacerbated by the pandemic and the shift to online interactions. However, children reported feeling lonely before the pandemic and again in 2021, indicating a need to facilitate socialization in settings where children spend time to promote their health and wellbeing.

### Limitations

This study employs a descriptive approach, utilising a self-report methodology to investigate children’s perceptions of their childhood. The present paper exclusively presents the findings derived from survey participants. However, it is important to note that the study does not delve into the underlying reasons behind children’s responses or explore the factors contributing to changes in their perspectives.

## Conclusion

In summary, the study indicates that the health and wellbeing of children had been deteriorating before the Covid-19 pandemic and has either stabilised or continued to decline following the pandemic. Thus, it cannot be assumed that the removal of restrictions and return to normal routines and school will automatically improve children’s health and wellbeing. Therefore, it is imperative to prioritise the development and implementation of effective and sustainable interventions, funding distribution and policy focus that address physical skills like swimming and cycling, promote confidence and autonomy in physical activity, and enhance overall wellbeing and socialization. It is also essential to recognize that the pandemic and its accompanying restrictions could exacerbate these issues, and broader political and societal challenges like Brexit and the “cost of living” crisis may further complicate matters.

## Data Availability

Data is available upon reasonable request. The HAPPEN Dataset is available to access via the SAIL Databank.

## Acknowledgements

This research was funded by the National Centre for Population Health and Wellbeing and ADR UK. The research team would like to thank all pupils, teachers and schools who took part in facilitating, administering, and taking part in the HAPPEN Survey and for their support.

## References

1. Daines CL, Hansen D, Novilla MLB, Crandall AA. Effects of positive and negative childhood experiences on adult family health. BMC Public Health. 2021;21:1–8.

2. Mental Health Foundation. Children and young people: Statistics. 2023.

3. Lloyd A, McKay RT, Furl N. Individuals with adverse childhood experiences explore less and underweight reward feedback. Proc Natl Acad Sci U S A. 2022;119:1–8.

4. Lebrun-Harris LA, Ghandour RM, Kogan MD, Warren MD. Five-Year Trends in US Children’s Health and Well-being, 2016-2020. JAMA Pediatr. 2022;176:2016–20.

5. Peytignet S, Marszalek K, Grimm F, Thorlby R, Wagstaff T. Children and young people’s mental health. Heal Found. 2022.

6. NHS Digital. Mental Health of Children and Young People in England 2022 - wave 3 follow up to the 2017 survey. 2022. https://digital.nhs.uk/data-and-information/publications/statistical/mental-health-of-children-and-young-people-in-england/2022-follow-up-to-the-2017-survey#highlights. Accessed 13 Feb 2023.

7. Department for Education. State of the Nation 2021: Children and young people’s wellbeing research report. 2022; February.

8. Welsh Government. Wellbeing of Wales 2022: Children and young people’s wellbeing. Cardiff; 2022. https://gov.wales/wellbeing-wales-2022-children-and-young-peoples-wellbeing f.

9. Basterfield L, Burn NL, Galna B, Batten H, Goffe L, Karoblyte G, et al. Changes in children’s physical fitness, BMI and health-related quality of life after the first 2020 COVID-19 lockdown in England: A longitudinal study. J Sports Sci. 2022;40:1088–96. doi:10.1080/02640414.2022.2047504.

10. Government W. Wellbeing of Future Generations Act (Wales) 2015. 2015.

11. Welsh Government. Wales: A play friendly country. Cardiff; 2014.

12. Viner RM, Russell SJ, Croker H, Packer J, Ward J, Stansfield C, et al. School closure and management practices during coronavirus outbreaks including COVID-19: a rapid systematic review. Lancet Child Adolesc Heal. 2020;4:397–404. doi:10.1016/S2352-4642(20)30095-X.

13. Viner R, Russell S, Saulle R, Croker H, Stansfield C, Packer J, et al. School Closures during Social Lockdown and Mental Health, Health Behaviors, and Well-being among Children and Adolescents during the First COVID-19 Wave: A Systematic Review. JAMA Pediatr. 2022;176:400–9.

14. James M, Marchant E, Defeyter MA, Woodside J V., Brophy S. Impact of School Closures on the Health and Well-Being of Primary School Children in Wales UK; A Routine Data Linkage Study Using the HAPPEN Survey (2018-2020). SSRN Electron J. 2021;:1–19.

15. BBC Children in Need. Understanding the impact of the Covid-19 Pandemic on children and young people. 2020; May. https://www.bbcchildreninneed.co.uk/wp-content/uploads/2020/11/CN1081-Impact-Report.pdf.

16. Fahy N, Hervey T, Dayan M, Flear M, Galsworthy MJ, Greer S, et al. Impact on the NHS and health of the UK’s trade and cooperation relationship with the EU, and beyond. Heal Econ Policy Law. 2022;17:471–96.

17. Goddard A. The cost of living crisis is another reminder that our health is shaped by our environment. BMJ. 2022;:35595270.

18. Mental Health Foundation. Mental health and the cost-of-living crisis report: another pandemic in the making? 2022.

19. Todd C, Christian D, Davies H, Rance J, Stratton G, Rapport F, et al. Headteachers’ prior beliefs on child health and their engagement in school based health interventions: A qualitative study. BMC Res Notes. 2015;8:1–10. doi:10.1186/s13104-015-1091-2.

20. Todd C, Chistian D, Tyler R, Stratton G, Brophy S. Developing HAPPEN (Health and Attainment of Pupils involved in a Primary Education Network): working in partnership to improve child health and education. Perspect Public Health. 2016;136:115–6.

21. StataCorp. Stata Statistical Software: Release 16. 2019;College St StataCorp LLC.

22. R Core Team. R: A language and environment for statistical computing. Vienna, Austria: R Foundation for Statistical Computing; 2021. https://www.r-project.org/.

23. Whitehead M. The definition of physical literacy. 2016. https://www.physical-literacy.org.uk/defining-physical-literacy/. Accessed 17 Jan 2020.

24. Invernizzi PL, Rigon M, Signorini G, Alberti G, Raiola G, Bosio A. Aquatic physical literacy: The effectiveness of applied pedagogy on parents’ and children’s perceptions of aquatic motor competence. Int J Environ Res Public Health. 2021;18.

25. Sport Wales. Sport Wales Welsh Government Free Swimming Initiative (FSI) – a new approach. 2018;:16–8.

26. Marzi I, Reimers AK. Children’s independent mobility: Current knowledge, future directions, and public health implications. Int J Environ Res Public Health. 2018;15.

27. Pozoukidou G, Chatziyiannaki Z. 15-minute city: Decomposing the new urban planning Eutopia. Sustain. 2021;13:1–25.

28. Department for Transport, England AT. Bikeability receives record £20 million government investment to improve access to cycle training. 2022. https://www.gov.uk/government/news/bikeability-receives-record-20-million-government-investment-to-improve-access-to-cycle-training. Accessed 13 Mar 2023.

29. Marchant E, Todd C, James M, Crick T, Dwyer R, Brophy S. Primary school staff perspectives of school closures due to COVID-19, experiences of schools reopening and recommendations for the future: A qualitative survey in Wales. PLoS One. 2021;16 12 December:1–24. doi:10.1371/journal.pone.0260396.

30. Marchant E, Todd C, James M, Crick T, Dwyer R, Kingdom U, et al. Primary school staff reflections on school closures due to COVID-19 and recommendations for the future: a national qualitative survey. 2020.

31. Parnham JC, Laverty AA, Majeed A, Vamos EP. Half of children entitled to free school meals did not have access to the scheme during COVID-19 lockdown in the UK. Public Health. 2020;187 March:161–4. doi:10.1016/j.puhe.2020.08.019.

32. Jacques-Avinõ C, López-Jiménez T, Medina-Perucha L, De Bont J, Goncąlves AQ, Duarte-Salles T, et al. Gender-based approach on the social impact and mental health in Spain during COVID-19 lockdown: A cross-sectional study. BMJ Open. 2020;10:1–10.

33. Waite P, Pearcey S, Shum A, Raw JAL, Patalay P, Creswell C. How did the mental health symptoms of children and adolescents change over early lockdown during the COVID-19 pandemic in the UK? JCPP Adv. 2021;1:1–10.

34. James M, Jones H, Baig A, Marchant E, Waites T, Todd C, et al. Factors influencing wellbeing in young people during COVID-19: A survey with 6291 young people in Wales. PLoS One. 2021;16 12 December:1–17. doi:10.1371/journal.pone.0260640.

35. Afshordi N. Children’s Inferences About Friendship and Shared Preferences Based on Reported Information. Child Dev. 2019;90:719–27.

